# Highly Accurate and Reproducible Diagnosis of Peanut Allergy Using Epitope Mapping

**DOI:** 10.1101/2020.06.19.20136002

**Authors:** Paul Kearney, Robert Getts, Clive Hayward, David Luta, Alex Porter, Marc Witmer, George du Toit, Gideon Lack, R. Sharon Chinthrajah, Stephen J Galli, Kari Nadeau, Galina Grishina, Mayte Suárez-Fariñas, Maria Suprun, Hugh A Sampson

## Abstract

**Background:** Misdiagnosis of peanut allergy is a significant clinical challenge. Here, a novel diagnostic blood-based test using a Bead-Based Epitope Assay (“peanut BBEA”) has been developed on the LEAP cohort and then independently validated on the CoFAR2 and POISED cohorts.

**Methods:** Development of the peanut BBEA followed the National Academy of Medicine’s established guidelines with discovery performed on 133 subjects from the non-interventional arm of the LEAP trial and an independent validation performed on 81 subjects from the CoFAR2 study and 84 subjects from the POISED study. All subject samples were analyzed using the BBEA methodology. The peanut BBEA test measures levels of two Ara h 2 epitopes and compares their combination to a pre=specified threshold. If the combination of the two epitope levels is at or below the threshold, then the subject is ruled “Not Allergic”, otherwise the subject is ruled “Allergic”.

All allergic diagnoses were OFC confirmed and subjects’ ages were 7-55 years.

**Results:** In validation on the CoFAR2 and POISED cohorts, the peanut BBEA test had a combined sensitivity, specificity, positive predictive value, negative predictive value, positive likelihood ratio, negative likelihood ratio and accuracy of 91%, 95%, 95%, 91%, 18.2, 0.09 and 93%, respectively.

**Conclusion:** The peanut BBEA test performance in validation demonstrated overall high accuracy and compared very favorably with existing diagnostic tests for peanut allergy including skin prick testing, peanut sIgE and peanut component testing.

## Introduction

Prevalence of peanut allergy among children in the United States is estimated to be ∼2% [1]. The gold standard for diagnosis of peanut allergy is the double-blind placebo-controlled food challenge (DBPCFC), however, the DBPCFC is time- and resource-consuming, not widely available, and a potentially risky procedure [2]. More commonly, diagnosis is determined using a combination of patient history, the skin prick test (SPT), and peanut specific IgE (sIgE) and peanut allergen component specific testing (e.g. Ara h 2) [3-6]. Heuristics for optimally using these tools, on their own or in combination, have been explored [4-5]. Nevertheless, diagnostic performance of these tools has fallen short both in terms of accuracy and level of evidence. In particular, it is estimated that the rate of peanut allergy “over-diagnosis” is over 60% [7]. More recently the Basophil Activation Test (BAT) has been assessed demonstrating higher accuracy than the SPT and Ara h 2 testing (used independently) but statistical superiority was not established [8]. Additionally, BAT testing has been associated with a 5-10% non-responsive rate to IgE-mediated stimulation [9] and requires fresh whole blood.

An ideal alternative diagnostic test for the oral food challenge would have the following attributes:

- Be relatively non-invasive, such as a blood-based test;
- Be rigorously developed and documented in accordance with national guidelines such as those established by the National Academy of Medicine [10];
- Be clinically validated on multiple independent and well-characterized cohorts;
- Be clinically validated on subjects whose allergy status has been largely confirmed by DBPCFC;
- Be clinically and analytically validated in a laboratory with documented standard operating procedures, and ideally, certified and accredited by an external regulatory group such as CLIA or CAP;
- Has diagnostic performance that is at least 90% accurate as compared to DBPCFC; and
- Statistically superior to established clinical tests for diagnosis.

These criteria ensure that the alternative diagnostic test is accurate with both a high level of clinical and analytical validation.

Here we present the discovery and clinical validation of a test for the diagnosis of peanut allergy that meets the criteria above. The diagnostic test, or peanut Bead-Based Epitope Assay (peanut BBEA), is a plasma-based test that utilizes the previously characterized BBEA for measuring IgE antibody binding to sequential peanut epitopes in a patient blood sample [11]. The assay enables characterization of the biological mechanism of IgE-mediated allergic response where two epitopes of a peanut allergen protein crosslink cell surface bound IgE molecules resulting in degranulation and initiation of an allergic response. Here we describe the application of this methodology in two independent clinical validations, both of which demonstrated sensitivity and specificity above 90% in comparison to DBPCFC, and accuracy significantly and statistically superior to established diagnostic tests such as the SPT, sIgE and peanut allergen component testing.

## Methods

### BBEA Assay

The BBEA assay was performed as previously described [11]. Briefly, sixty-four clinically-relevant peptides belonging to Ara h 1 (n=34), Ara h 2 (n=16), and Ara h 3 (n=14) were synthesized (CS Bio, Menlo Park, CA, USA), coupled to LumAvidin beads (Luminex Corporation, Austin, TX, USA) and stored in PBS-TBN buffer (1xPBS + 0.02%Tween20 + 0.1%BSA). A master mix of peptide-coupled beads was made in PBS-TBN buffer and 100uL of the bead master mix was added to 96-well filter plates. After washing the beads, 100μL of the subject’s plasma at 1:10 dilution was added to the wells. The plates were incubated on a shaker for 2 hours at 300rpm at room temperature. Excess plasma was then removed, and the plate was washed. 50μL/well of mouse anti-human IgE-PE (Thermo-Pierce Antibodies, Clone BE5, diluted 1:50 in PBS-TBN), secondary antibody was added, and plates were incubated for 30 minutes. After a final wash, 100μL of PBS-TBN buffer was added to each well to re-suspend the beads, which were then transferred to fixed-bottom 96-well reading plates, and quantified as Median Fluorescence Intensity (MFI) on the Luminex 200 instrument (Luminex^®^ 100/200™ System, Luminex Corporation, Austin, TX, USA).

### Discovery

Discovery of the peanut BBEA was performed on 133 subjects (31 allergic, 102 non-allergic) from the avoidance arm of the LEAP study (NCT00329784) where all diagnoses were confirmed by DBPCFC. Plasma samples were obtained at approximate years 2 and 5 for each subject. These samples were analyzed using the BBEA methodology to obtain the IgE epitope reactivity levels.

The best diagnostic pair of epitopes was determined at year 5 by exhaustive search over all epitopes pairs and integrated into a minimum detectability model where the lower limit of detection for each epitope was determined. Linear regression was used to combined pairs of epitopes into models. The optimal model was then assessed at year 2 to ensure consistency. The decision threshold that maximized accuracy was determined and documented (fully locked-down prior to validation).

### Validation

Validation of the locked-down peanut BBEA was performed on two independent cohorts of subjects from CoFAR2 (NCT00356174) and POISED (NCT02103270). In both studies all subjects evaluated had their allergy status confirmed by DBPCFC.

Plasma samples were obtained for 82 subjects (23 allergic, 59 non-allergic) from the CoFAR2 study at years 2 and 5 for each subject. Additional plasma samples were obtained for 42 allergic subjects from the POISED study and 42 age-matched non-allergic subjects collected under a separate IRB approved protocol. POISED subjects had ages ranging from 7 to 55 years. All samples were analyzed using the BBEA methodology to obtain the IgE epitope reactivity levels.

### Data Analysis

For all three cohorts, subjects were randomized across plates and assayed in triplicate [13]. Raw MFI data were log-normalized, backgrounds subtracted, triplicates combined into a single value using the median function and plate-normalized using a plate standard sample.

Performance of the peanut BBEA was determined by constructing the confusion matrix of DBPCFC vs. peanut BBEA allergy classifications and then deriving sensitivity, specificity as well as other diagnostic metrics. The confusion matrix is a 2×2 matrix with a tally of the true positives, false positives, false negatives and true negatives. Statistical significance was determined by Fisher’s exact test. All data analyses were performed using Matlab R2019b.

## Results

The demographic profiles of the three cohorts (LEAP, CoFAR2 and POISED) are presented in Table 1. Note that the LEAP cohort was used for discovery whereas CoFAR2 and POISED were used to validate the algorithm derived from LEAP. The CoFAR2 validation cohort covered ages from 1 to over 5 years of age whereas POISED covered the age group from above 5 to 55 years of age. Allergic subject diagnosis was confirmed by DBPCFC. For LEAP and CoFAR2, DBPCFCs were performed at the year 5 visit.

**Table 1:** Demographic profiles of the three cohorts participating in the discovery and validation of the peanut BBEA test. Each cell contains the median and range of values. Ages for LEAP and CoFAR2 are reported for subjects at the time of enrollment into the longitudinal studies. Non-allergic controls for the POISED cohort did not have sIgE performed.

|  | LEAP |  | CoFAR2 |  | POISED |  |
| --- | --- | --- | --- | --- | --- | --- |
|  | Allergic | Non-Allergic | Allergic | Non-Allergic | Allergic | Non-Allergic |
| n | 31 | 102 | 23 | 59 | 42 | 42 |
| Enrollment Age (years) | 0.66<br>(0.37 - 0.91) | 0.68<br>(0.40 - 0.91) | 0.87<br>(0.37 - 1.23) | 0.81<br>(0.3 - 1.25) | 11<br>(7 - 49) | 10<br>(7 - 55) |
| slgE (kU<sub>A</sub>/L) | 0.39<br>(0.01 - 79.50) | 0.04<br>(0.01 - 87.70) | 1.46<br>(0.00 - 24.45) | 0.53<br>(0.00 - 60.32) | 37.4<br>(0.41 - 869) | N/A |

### Discovery

Two principles drive the derivation of the peanut BBEA algorithm:

- SPT and sIgE are effective for ruling-in and ruling-out peanut allergy at high and low values, respectively.
- IgE-mediated allergic reactions require two epitopes from the same peanut allergen to crosslink IgE.

Previous work has established that sufficiently high and low values for the SPT and sIgE test are needed to rule-in and rule-out peanut allergy [4-5]. Consequently, logistic regression was applied to all pairs of epitopes from the same peanut allergen (Ara h 1, Ara h 2 or Ara h 3) to determine which were most highly associated with DBPCFC allergy status for subjects with non-extreme sIgE/SPT values. Furthermore, it was required that these epitopes were reliably detected in all allergic subjects. In the LEAP cohort (5-year visit) the epitope pair from the same peanut allergen with the highest Area Under the Curve (AUC) and reliably detected were Ara h 2.008 and Ara h 2.019 (AUC = 74%). This pair of epitopes was assessed at the 2 year visit and had consistent performance (AUC = 73%). Additional epitopes did not improve the performance significantly, and so, the algorithm was restricted to these two epitopes to avoid overfitting. Combining the two principles above resulted in the following algorithm:

If SPT <= 3mm or sIgE <=0.1 kU_A_/L then “Not Allergic.”

If SPT >= 18mm or sIgE >= 18 kU_A_/L then “Allergic.”

If Ara h2.008 + Ara h2.019/20 <= 0.20 then “Not-Allergic”, otherwise “Allergic.”

In this specification, low and high values (e.g. 3mm and 18mm for the SPT) were identified based on previous work [14]. The analytical lower limit of detection was determined (using the Clinical and Laboratory Standards Institute (CLSI) guidelines) for epitopes Ara h 2.008 and Ara h

2.019 and resulted in the detection threshold (0.20). Effectively, if IgE reactivity to either of these two epitopes is sufficiently high, then the result is positive. This algorithm was documented and locked-down for evaluation on validation cohorts.

### Validation

With the laboratory procedures and peanut BBEA algorithm locked-down, the validation cohorts (CoFAR2 and POISED) were analyzed. In order to confirm that Ara h 2.008 and Ara h 2.019 reproduced as the two highest performing epitopes from the same peanut allergen, the AUC was determined for the CoFAR2 and POISED cohorts for all epitope pairs included in the BBEA panel. Figure 1 depicts the pairwise epitope AUC plot for all pairs of epitopes for the LEAP, CoFAR2 and POISED cohorts. This figure demonstrates which epitope pairs are synergistic, as well as the reproducibility across discovery and validation cohorts. Pairs of epitopes within the same allergen (Ara h 1, Ara h 2 and Ara h 3) with the highest AUC are within Ara h 2. Specifically, Ara h 2.008 and Ara h 2.019 have the highest combined AUC in both CoFAR2 and POISED, reproducing the Discovery observations in LEAP.

**Figure 1.**
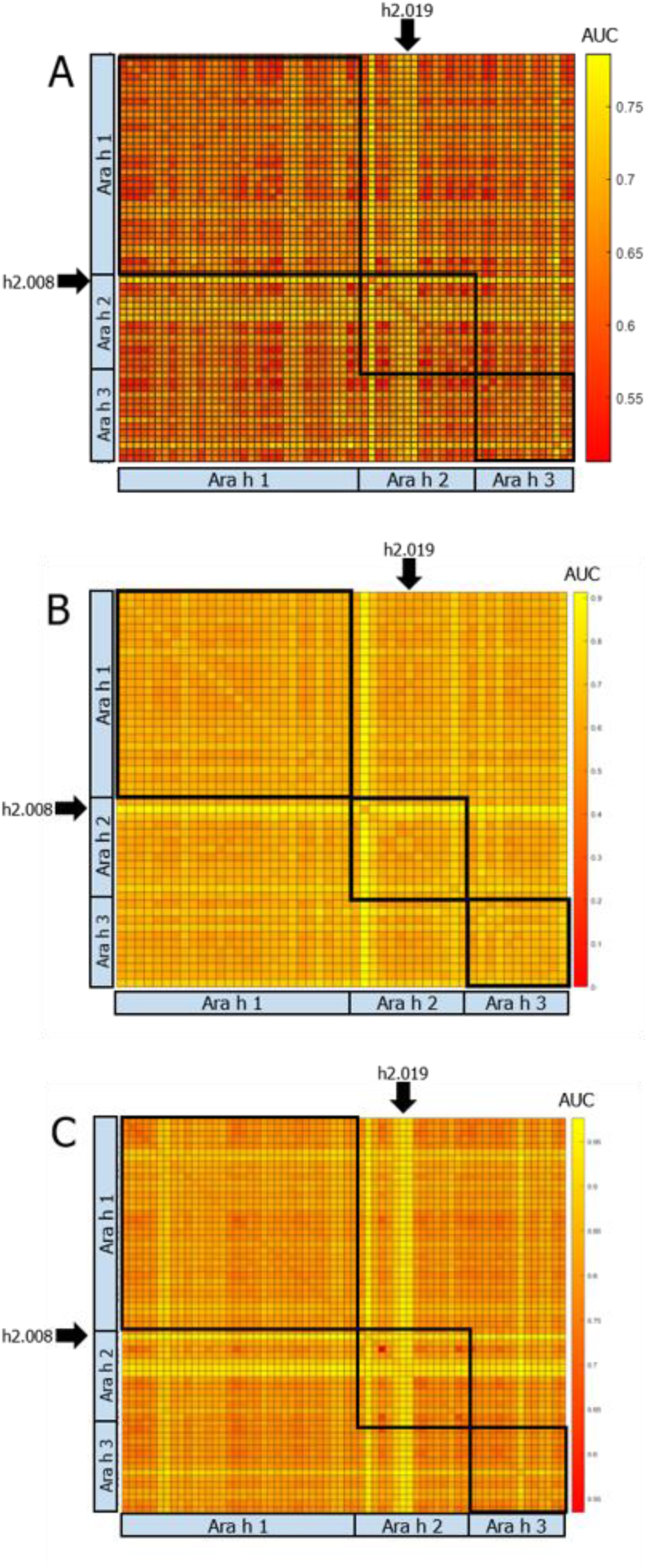
AUC heatmaps for all pairs of peanut allergen epitopes for LEAP (A), CoFAR2 (B) and POISED (C). Each heatmap pixel represents the AUC for the logistic regression classifier built over the pair of associated epitopes for classifying allergy status. Colors closer to yellow indicate a higher AUC.

To quantify the reproducibility of the peanut BBEA locked-down algorithm, confusion matrices comparing the DBPCFC classifications to the peanut BBEA classifications were generated for CoFAR2 and POISED. From the confusion matrices performance, Table 2 was derived. The accuracy of the peanut BBEA was highly significant by Fisher’s exact test (p-value <0.001). Table 2 also compares the performance of the peanut BBEA test to established tests (SPT, sIgE, component proteins). Although the Peanut BBEA method was locked-down before performance was assessed on CoFAR2 and POISED, the performance of established tests were assessed with both pre-specified thresholds and with optimization on CoFAR2. This provided the most conservative comparison of the Peanut BBEA method.

**Table 2:** Performance of Peanut BBEA in Validation on the CoFAR2 and POISED cohorts. Performance of SPT, sIgE and to peanut and its component proteins on the CoFAR2 cohort is presented for comparison. Commonly used thresholds selected to optimize sensitivity for the SPT, sIgE, Ara h 1, Ara h 2 and Ara h 2 tests, respectively, are 3mm, 0.1kU_A_/L, 0.30kU_A_/L, 0.30kU_A_/L and 0.30kU_A_/L. Performance at these thresholds are displayed for rows demarked with *. Thresholds optimizing accuracy were also determined for each serological tests and performance displayed (rows demarked with **). PPV, NPV, FPR, FNR, LR+ and LR- are positive predictive value, negative predictive value, false positive rate, false negative rate, positive likelihood ratio and negative likelihood ratio, respectively.

| Test | Sens. | Spec. | PPV | NPV | FPR | FNR | LR+ | LR- | Accuracy (CIs) |
| --- | --- | --- | --- | --- | --- | --- | --- | --- | --- |
| **Peanut BBEA (CoFAR2)** | 91% | 92% | 92% | 91% | 8% | 9% | 11.4 | 0.1 | 92% (91%-93%) |
| **Peanut BBEA (POISED)** | 93% | 98% | 98% | 93% | 2% | 7% | 46.5 | 0.07 | 95% (93%-97%) |
| **Peanut BBEA (Combined)** | 91% | 95% | 95% | 91% | 5% | 9% | 18.2 | 0.09 | 93% (91%-94%) |
| **SPT\* (CoFAR2)** | 96% | 61% | 71% | 93% | 39% | 4% | 2.5 | 0.07 | 71% (70%-77%) |
| **slgE\* (CoFAR2)** | 91% | 42% | 61% | 83% | 48% | 9% | 1.6 | 0.2 | 56% (53%-59%) |
| **Ara h 1\* (CoFAR2)** | 78% | 40% | 57% | 65% | 60% | 22% | 1.3 | 0.6 | 51% (48%-56%) |
| **Ara h 2\* (CoFAR2)** | 83% | 39% | 57% | 69% | 61% | 17% | 1.4 | 0.4 | 51% (50%-58%) |
| **Ara h 3\* (CoFAR2)** | 91% | 9% | 50% | 50% | 91% | 9% | 1 | 1 | 29% (27%-35%) |
| **slgE\*\* (CoFAR2)** | 61% | 85% | 61% | 85% | 15% | 39% | 4.0 | .46 | 78% (76%-81%) |
| **Ara h 1\*\* (CoFAR2)** | 43% | 86% | 56% | 79% | 14% | 57% | 3.1 | .66 | 72% (69%-76%) |
| **Ara h 2\*\* (CoFAR2)** | 78% | 79% | 60% | 90% | 21% | 22% | 3.7 | .28 | 77% (75%-81%) |
| **Ara h 3\*\* (CoFAR2)** | 83% | 32% | 33% | 82% | 68% | 17% | 1.2 | .55 | 45% (42%- 49%) |

Confidence intervals (95%) for accuracy were established using bootstrapping for Peanut BBEA and other tests on CoFAR2 [12]. As commonly used thresholds for the serological tests did not yield good performance characteristics, the threshold for each serological test optimizing accuracy was determined on CoFAR2 and performance re-assessed. It should be noted that the threshold for the peanut BBEA test was pre-defined and not optimized for the CoFAR2 or POISED cohorts. Regardless of whether or not the serological tests were optimized, the peanut BBEA test demonstrates statistically significant superior results as demonstrated by non-overlapping confidence intervals on the accuracy metric.

## Discussion

The need for accurate diagnosis of peanut allergy, an alternative for oral food challenges in clinical practice, is apparent more than ever with the onset of immunotherapies for peanut allergy. This underscores the importance of a diagnostic that is properly validated, analytically and clinically, with multiple cohorts and in the context of a regulated, qualified laboratory to ensure reproducible, reliable results.

It has been established in multiple cohorts that the peanut BBEA has accuracy, as measured against DBPCFC, of 93% for the diagnosis of peanut allergy. This compares very favorably to SPT, sIgE and peanut allergen component tests. Not only is the peanut BBEA substantially more accurate, but the false positive rate (FPR) is more than 7-fold lower than all other diagnostic tests.

An unexpected discovery presented here is the immunodominance of Ara h 2 epitope h2.008. Referring to Figure 1, it is apparent that consistently across three cohorts (LEAP, CoFAR2, POISED) h2.008 produces ahigh diagnostic AUC when paired with any other epitope from Ara h 2 and in particular h2.019. This suggests that IgE-mediated reactions predominantly occur when there is IgE reactivity with h2.008. While it is not surprising that Ara h 2 epitopes are dominant based on previous studies [15, 16, 17], the observation that >93% of allergic responses can be linked to just one epitope, h2.008, is intriguing. This has both therapeutic and diagnostic implications and requires further investigation.

## Data Availability

Data supporting the manuscript is available by contacting.

## Acknowledgements

The authors would like to acknowledge the contributions of the CoFAR investigators (Scott Sicherer, Robert Wood, Wesley Burks, Stacie Jones, and Donald Leung); POISED investigator Mindy Tsai; and funding from the Sean N. Parker Center for Allergy and Asthma Research at Stanford University.

